# Measuring College Students’ with Disabilities Attitudes Toward Taking COVID-19 Vaccines

**DOI:** 10.1101/2021.11.04.21265923

**Authors:** Z.W. Taylor, Chelseaia Charran

## Abstract

This survey explores attitudes of 245 currently enrolled college students with disabilities regarding their comfort taking a COVID-19 vaccine. Results suggest most college students with disabilities are willing to take a COVID-19 vaccine if their institution requires it to return to campus in subsequent semesters. However, many students with disabilities would not feel comfortable with a vaccine mandate mid-semester and would consider withdrawing, especially among older students with disabilities and first-generation college students with disabilities. Implications for postsecondary policy and leadership are addressed.

## How Do College Students with Disabilities Feel About Taking COVID-19 Vaccines?

After March 11th, 2020, when the World Health Organization (WHO) declared COVID-19 (coronavirus) a global pandemic (World Health Organization, 2020), colleges and universities briskly moved students, faculty, and staff online in an abundance of safety and caution. This move to online learning was particularly burdensome, both in terms of human and financial capital, for institutions with large on-campus populations. Moving students online also removed them from residence halls and student affairs related activities on campus, many of which employ thousands of higher education professionals across the United States and the world (Hubler, 2020). Additionally, on campus activities, such as becoming involved in student organizations and socializing with classmates in true in person settings have been proven to increase retention and graduation rates, compounding the difficulty of a shift to online learning (Bawa, 2016). Perhaps more so than any other student population, students with disabilities may have been most marginalized by this rapid move online, as students with disabilities often reported that institutions and their practitioners were not ready to support them in online spaces and many disability services struggled to deliver services in a timely manner (AHEAD, 2021).

Given the many hurdles presented by COVID-19, many institutions of higher education are exploring how to reopen their doors in subsequent semesters. Reopening would not only bring students back to campus but also bring faculty and staff back to campus and re-employ thousands of laid off or furloughed workers across the United States and the World (Hubler, 2020). However, no research has explored how students with disabilities view COVID-19 vaccines and whether these students feel safe taking a vaccine that may negatively interact with any one of their disabilities. As has been well-documented across dozens of news outlets, the development of a viable, safe, and effective COVID-19 vaccine became a reality in early 2021, as both Pfizer and Moderna mRNA vaccines cleared emergency authorization and were available to certain populations (Centers for Disease Control and Prevention, 2021). The public availability of vaccines in early 2021 was often limited to high-risk populations including those who are 65 and older or those who were essential workers working in healthcare, such as hospital and clinic workers (Centers for Disease Control and Prevention, 2021). Furthermore, there is an emerging debate as to whether colleges and universities can mandate vaccines, especially for at-risk populations such as students with disabilities (Gostin et al., 2021) or students with Constitutional grounds for vaccine waivers through claims to religious freedom (Reiss & DiPaolo, 2021).. Yet, little data has emerged that has focused on how COVID-19 vaccines affect people with disabilities differently than people without disabilities and whether these people feel comfortable taking COVID-19 vaccines as part of plans to physically return to postsecondary campuses.

As a result, as COVID-19 vaccines becomes more widely available, colleges and universities, especially those with robust on campus living facilities and residence halls, maybe face difficult questions as they relate to college students with disabilities:

1. How comfortable do college students with disabilities feel taking a COVID-19 vaccine?
2. Would college students with disabilities take a COVID-19 if it were required by their institution to return to campus?
3. Mid-semester, would students with disabilities withdraw from classes if their institution required a COVID-19 vaccine to stay on campus?

These are the questions that this brief answers through a survey of 237 currently-enrolled postsecondary students in the United States at all three levels: two-year students, four-year students, and graduate students. Results suggest many students with disabilities are willing to take a COVID-19 vaccine if their institution required it to return to on-campus learning, however some students may be wary of returning to campus without a vaccine only to be told mid-semester that a vaccine is required to stay on-campus. The research team will close the study by addressing implications for policy, research, and practice surrounding COVID-19 vaccinations and the health and safety of college students with disabilities.

## Methods

### Data Collection

Data for this survey were gathered in January 2021 when public availability of COVID-19 vaccines became clearer through official communication from the Centers for Disease Control and the World Health Organization (Centers for Disease Control and Prevention, 2021). The research team employed Amazon Mechanical Turk (MTurk) to survey students with disabilities currently enrolled at institutions of higher education. MTurk has been found to be a unique and robust source of human intelligence services, including survey completion in educational contexts (Follmer et al., 2017). Several recent studies in education focused on financial aid jargon (Taylor & Bicak, 2019) and computer science education (Hellas et al., 2020) have used MTurk to answer research questions that require a large, nationally representative dataset, akin to the study at hand related to postsecondary student attitudes toward COVID-19 vaccinations.

The survey asked for a student’s college enrollment status (yes or no) and a student’s dis/ability status (yes, has a disability or no, does not). Both questions required affirmative answers before a respondent could proceed. Then, the respondent was asked for birth year, race, gender, first generation in college status (defined as neither parent earning any level of postsecondary credential), educational level (two-year, four-year, or graduate), enrollment status (part- or full-time), and current mode of education (on-campus, online, or hybrid). Survey respondents came from nearly all fifty United States with the exception of North Dakota, South Dakota, Wyoming, and Puerto Rico. Respondents from California (12% of sample), Texas (10% of sample) of sample, New York (10% of sample), and Florida (9% of sample) were overrepresented in the overall survey respondent pool. A description of the respondents in this study’s survey can be found in Table 1 below:

**Table 1.**
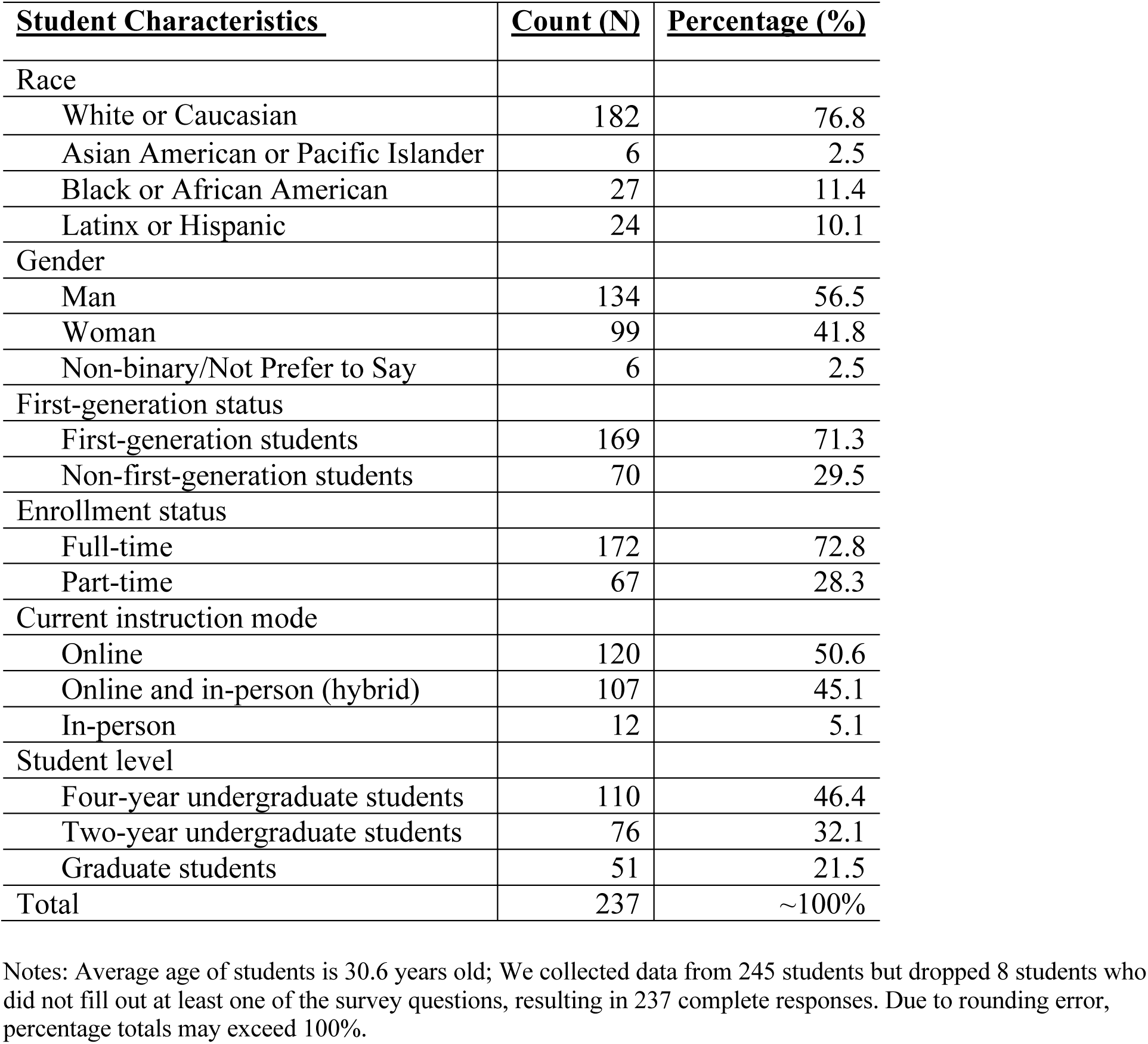
Descriptive statistics of survey sample and responses (n=237)

| **<u>Student Characteristics</u>** | **<u>Count (N)</u>** | **<u>Percentage (%)</u>** |
| --- | --- | --- |
| Race |  |  |
| White or Caucasian | 182 | 76.8 |
| Asian American or Pacific Islander | 6 | 2.5 |
| Black or African American | 27 | 11.4 |
| Latinx or Hispanic | 24 | 10.1 |
| Gender |  |  |
| Man | 134 | 56.5 |
| Woman | 99 | 41.8 |
| Non-binary/Not Prefer to Say | 6 | 2.5 |
| First-generation status |  |  |
| First-generation students | 169 | 71.3 |
| Non-first-generation students | 70 | 29.5 |
| Enrollment status |  |  |
| Full-time | 172 | 72.8 |
| Part-time | 67 | 28.3 |
| Current instruction mode |  |  |
| Online | 120 | 50.6 |
| Online and in-person (hybrid) | 107 | 45.1 |
| In-person | 12 | 5.1 |
| Student level |  |  |
| Four-year undergraduate students | 110 | 46.4 |
| Two-year undergraduate students | 76 | 32.1 |
| Graduate students | 51 | 21.5 |
| Total | 237 | ~100% |

Notes: Average age of students is 30.6 years old; We collected data from 245 students but dropped 8 students who did not fill out at least one of the survey questions, resulting in 237 complete responses. Due to rounding error, percentage totals may exceed 100%.

Then, students were asked to report their level of comfort regarding taking a COVID-19 vaccine on a seven-point Likert scale, followed by three follow-up questions:

1. What is your level of comfort taking a COVID-19 vaccine? (1=extremely comfortable, 7=extremely uncomfortable)
2. Would you take a COVID-19 vaccine if your school REQUIRED it for you to return to campus? (Yes/No)
3. Mid-semester, would you withdraw from classes at your school if your school REQUIRED a COVID-19 vaccine to stay on campus? (Yes/No)

### Data Analysis

Given the outcome variables of this study’s research question and only one year or set of observations in the data, the team decided upon an OLS regression model to predict vaccine comfort (1-7 scale) and logistic regression models (taking a vaccine, yes or no and withdrawing after a vaccine mandate, yes or no) to analyze this study’s data. As a result, the team transformed string variables into numerical data and assigned the largest population across all demographics as the control group for each regression (White students, not first generation students, students attending four-year institutions, etc.). Model 1, 2 and 3 each contain different numbers of observations as several survey responses perfectly predicted our outcome variables; thus, these students were removed from each Model and overall observations can be found in Table 3 across all three regression Models.

### Limitations

As with any survey study, this study is limited primarily by the reliability and validity of the survey data. This study gathered data from MTurk and participants who self-reported their college enrollment status and their dis/ability status. Moreover, this study is also limited by its temporal nature, meaning that students’ with disabilities attitudes toward COVID-19 vaccines may drastically change over time as vaccine efficacy is reported and vaccines become safer and more available to the general public and the disability community (Gostin et al., 2021; Reiss & DiPaolo, 2021). In addition, as institutions of higher education release their reopening plans for full on-campus immersion in the 2021 and 2022 academic years, college student attitudes towards taking a COVID-19 vaccine may also change due to idiosyncratic institutional planning. Yet, the strengths of this study is its sample size (n=237), rendering it robust for quantitative analysis and generalizability, while also reporting timely and critical data for institutions of higher education: For these reasons, the research team feels the study’s strengths outweigh its limitations.

## Results

Descriptive statistics of survey responses can be found in Table 2 below:

**Table 2.** College students with disabilities comfort with COVID-19 vaccines (n=237)

| How comfortable do you feel taking a COVID-19 vaccine? | **<u>Count</u>**<br>**<u>(N)</u>** | **<u>Percentage</u>**<br>**<u>(%)</u>** |
| --- | --- | --- |
| Extremely Comfortable | 72 | 30.4 |
| Moderately Comfortable | 60 | 25.3 |
| Slightly Comfortable | 27 | 11.4 |
| Neutral | 16 | 6.8 |
| Slightly Uncomfortable | 27 | 11.4 |
| Moderately Uncomfortable | 6 | 2.5 |
| Extremely Uncomfortable | 31 | 13.1 |
| Would you take a COVID-19 vaccine if your institution<br>REQUIRED it for you to return to campus? |  |  |
| Yes | 180 | 75.9 |
| No | 59 | 24.9 |
| Mid-semester, would you withdraw from classes at your institution<br>if your institution REQUIRED a COVID-19 vaccine to stay on<br>campus? |  |  |
| Yes | 157 | 66.2 |
| No | 82 | 34.6 |
| Total | 237 | 100% |

Overall, 30.4% of students with disabilities felt “extremely comfortable” taking a COVID-19 vaccine, while at least 25% of these students expressed at least slight discomfort in taking a vaccine. Additionally, over 75% of students with disabilities would take a COVID-19 vaccine if they needed one to physically return to campus in Fall 2021. Finally, 66.2% of students with disabilities reported that they would withdraw from classes mid-semester if their institution suddenly required a COVID-19 vaccine to stay on campus. Results from the latter two questions were surprising, as over 75% of respondents reported being willing to take a required vaccine to return to campus, yet 66.2% of respondents reported that they would withdraw if this vaccine requirement occurred mid-semester. Here, future research may be needed into how students with disabilities perceive vaccine mandates and whether the stress of seeking and receiving a vaccine may outweigh the educational benefits of on-campus learning and interaction.

Regression analyses predicting college students’ with disabilities levels of comfort with COVID-19 vaccines (OLS regression; Model 1), attitudes towards vaccine mandates to return to campus (logistic regression; Model 2), and likelihood of withdrawing mid-semester if an institution mandates COVID-19 vaccines (logistic regression; Model 3) can be found in Table 3 below:

**Table 3.**
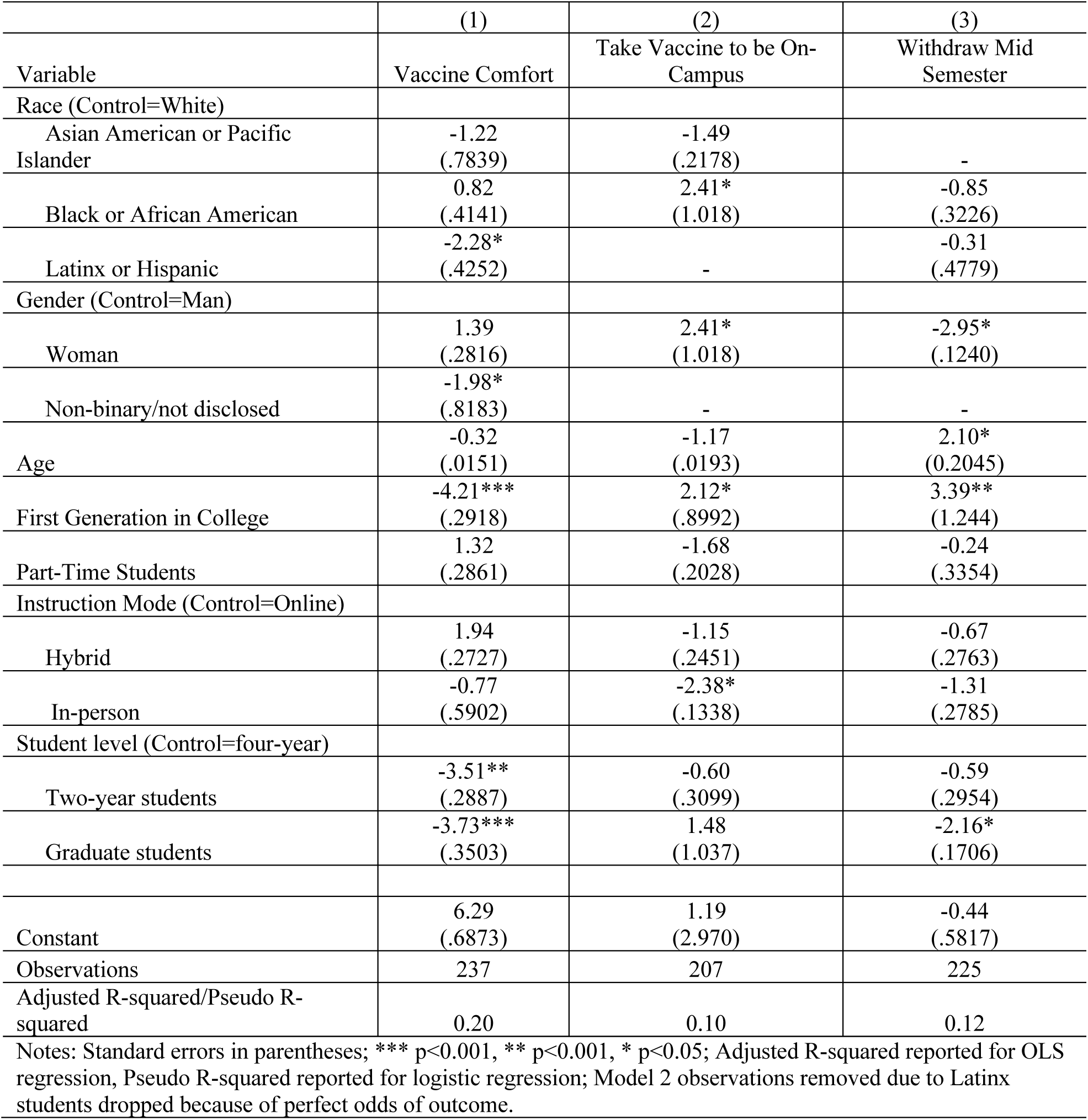
Regression analyses predicting college students’ COVID-19 vaccine attitudes (n=237)

Results in Table 3 and Model 1 suggest Latinx students with disabilities felt more comfortable with taking a COVID-19 vaccine (p < 0.05) when compared to students from different racial and ethnic backgrounds, controlling for other demographic characteristics. Similarly, non-binary identifying students (p < 0.05), first generation students (p < 0.00), community college students (p < 0.01), and graduate students (p < 0.00) felt more comfortable taking COVID-19 vaccines than their counterparts after controlling for other demographic characteristics. Attendance status (hybrid or in-person) and course load (full versus part time) was not predictive of COVID-19 vaccine comfort.

Results of the logistic regression in Model 2 suggest Black or African American students (p < 0.05), Latinx students (ideal model fit), women students (p < 0.05), and students attending courses in-person (p < 0.05) were more likely to take a COVID-19 vaccine to attend courses in-person/on-campus. Other demographic characteristics were not predictive of a students’ with a disabilities willingness to take a COVID-19 vaccine to return to in-person, on-campus learning. Moreover, results of the logistic regression in Model 3 suggest women students (p < 0.05) and graduate students (p < 0.05) are not as likely to withdraw mid-semester if their institution required COVID-19 vaccination, whereas older students (p < 0.05) and first generation students (p < 0.01) are more likely to withdraw mid-semester if their institution required COVID-19 vaccination compared to counterparts after controlling for other demographic characteristics. Collectively, these results suggest diverse attitudes toward COVID-19 vaccine mandates held by students with disabilities from a variety of intersectional identities.

## Discussion and Implications for Student Affairs Leadership and Campus Policy

Central to the conversation about universities’ Fall 2021 and beyond opening plans has focused on vaccine distribution and whether institutions felt it necessary to mandate COVID-19 vaccines to return to on-campus learning. However, data in this study suggest that students with disabilities who hold certain intersectional identities may not feel comfortable taking a COVID-19 vaccine for real, potentially life-threatening reasons given how COVID-19 vaccines may produce adverse effects or interactions with disability-related medications. For example, many students with disabilities may feel comfortable taking a COVID-19 vaccine to return to campus (∼75% of respondents, Table 2), yet many students indicated that they may withdraw from campus mid-semester if their institution mandated vaccines mid-semester (∼66% of respondents, Table 2). Here, institutions must understand that students with disabilities may require either extra time to receive a vaccine or may not feel comfortable recovering from a COVID-19 vaccine mid-semester when they are on-campus and potentially away from their family and primary medical provider. As a result, the timing of vaccine mandates may be key to fostering a sense of comfort and willingness to take a COVID-19 vaccine by students with disabilities. Furthermore, if there is a change of vaccine mandates mid-semester, universities should consider offering online learning for students with disabilities who are not comfortable taking the vaccine right away, so they do not have to withdraw from their classes.

Moreover, regression analyses in Table 3 suggest that certain students with disabilities may hold intersectional identities that could influence their vaccine comfort, their willingness to take a mandated vaccine, and their willingness to take a vaccine mid-semester when they are on-campus and potentially away from their support systems. Notably, women with disabilities expressed a strong willingness to take a COVID-19 vaccine to return to campus, whether that mandate was made prior to the semester or mid-semester. However, first generation students with disabilities expressed a willingness to take a COVID-19 prior to the semester but not mid-semester. These attitudes may be attributed to a variety of factors related to their intersectional identities and the hurdles they may face in procuring appropriate medical care while on-campus and away from home. From here, institutional leaders and student affairs professionals must be cognizant of the potentially minoritizing effect of COVID-19 vaccination policies as sections of plans to return to on-campus learning--these mandates may influence more first-generation students with disabilities off-campus, which would continue to perpetuate a cycle of discrimination that these students have already faced in non-pandemic times.

Ultimately, many institutional communications have focused on the necessity for vaccines to be widely available and accessible to the staff, students, and faculty that make up university communities. While COVID-19 variants and community surges are to be expected in the foreseeable future, the progress of vaccines and the potential to inoculate entire student bodies and personnel presents promise. However, vaccine shortages, modes of delivery, and individual willingness to become vaccinated remain an ongoing hurdle for higher education officials. What is not reported on, however, is how students with disabilities feel toward COVID-19 vaccination mandates. Particularly, that these vaccination mandates may not be safe for them while they are away from home and their primary care, or these mandates may not be safe given the type and intersectional nature of a student’s disability.

Ultimately, it is critical for postsecondary institutions to communicate COVID-19 vaccination mandates clearly and concisely to all students, especially students with disabilities, in a timely manner, to ensure that vaccine procedures are safe and can be conducted in ways that do not further marginalize this population. If colleges and universities across the United States want to safely open campuses in Fall 2021 and beyond, vaccine mandates should be carefully considered and implemented, as taking a vaccine for any purpose may indeed be a life-or-death decision for a college student with a disability.

## Data Availability

All data produced in the present study are available upon reasonable request to the authors

